# Performance of a Large Language Model in BI-RADS Classification of Ultrasound Based Breast Lesions

**DOI:** 10.1101/2025.09.28.25336860

**Authors:** Kathryn Pillai, Fauzia Nausheen

**Author notes:** **CORRESPONDING AUTHOR** Kathryn Pillai, **Email:**, **Address:** Department of Medical Education, California University of Science and Medicine, School of Medicine. 1501 Violet St, Colton, CA, USA. **FUNDING:** No funding was provided.

## Abstract

**Aims:** Given the advent of large language models (LLMs), the number of potential applications using artificial intelligence technologies in radiology has rapidly increased. Recently, several studies have evaluated the accuracy and quality of LLMs to characterize CT and MRI scans. Yet, to our knowledge, there have been few studies that have reported the utility of these models in generating BI-RADS assessment categories.

**Methods:** A breast ultrasound dataset including 256 images from 256 patients manually interpreted and labeled by radiologists according to BI-RADS features and lexicon was used for evaluating Gemini 2.0 Flash. We prompted the model to assess images in individual context windows and tested it with two variations of the original prompt (*n* = 3). Statistical analyses were then performed comparing the abilities of the model to the ground truth. The receiver operating characteristic-area under the curve (ROC-AUC) analysis was then calculated for each classification type from individual replicates.

**Results:** We found that the overall accuracy of Gemini 2.0 was 19.01% in predicting the BI-RADS classification of the breast lesions, and those of each category did not significantly differ from one another. From the ROC-AUC analysis, all category scores ranged from 0.5-0.6, and found that the model performed slightly better at categorizing benign lesions (1-4a), while those of greater probability of malignancy were akin to random chance (4b-5). Furthermore, we found that among incorrect predictions, the model was generally within 1-2 categories away from the true classification, demonstrating a low precision unreliable for realistic clinical usage.

**Conclusions:** This work highlights the current limitations of artificial intelligence models in classifying clinical images, and further development is required in these technologies before translation into the clinical setting. To our knowledge, this is the first study to report the capabilities of LLMs in performing BI-RADS classification of breast lesions with replicates.

## 1. Introduction

Large language models (LLMs) are artificial intelligence systems that have been trained upon billions of text-based content, and are able to engage in natural language processing tasks without having prior training [1-3]. In the past two years, LLMs such as OpenAI’s ChatGPT have shown capabilities to engage in the clinical decision-making and medical education as it had scored within passing range of the United States Medical Licensing Exam (USMLE) Step 1, Step 2CK, and 3 [4]. Since its reporting in early 2023, it has inspired countless studies evaluating the performance of LLMs in disease diagnostics [5-6], treatment planning [6-8], and in the healthcare setting [9]. From all medical specialities, the advent of these technologies has been most applicable in radiology applications [10]. This is well described by the introduction of multimodal LLMs that are able to evaluate both images and text-based contents. In the field of radiology, LLMs with multimodal capabilities have been used for generating reports from CT and MRI scans, assisting in differential diagnoses, EHR summarization, and patient generation [11]. Yet, there have been few studies that have reported the capabilities of these models in interpreting and classifying mammograms, ultrasounds, and MRI images. The Breast Imaging Report and Data System (BI-RADS) developed by the American College of Radiology is the foundational method for screening these images as it considers the densities and masses, micro/microcalcifications, architectural distortions, special cases (ductal ectasia, intramammary lymph node, or focal asymmetric density), and other associated findings (skin or nipple retraction, skin thickening, cutaneous lesions, axillary lymph nodes) [12]. BI-RADS classifies images in 6 categories, including 0 (incomplete), 1 (normal), 2 (benign), 3 (probably benign), 4a (suspicious abnormality but very low probability of malignancy), 4b (suspicious abnormality with intermediate probability of malignancy), 4c (suspicious abnormality with non-typical risk of malignancy), 5 (suggestive of malignancy), and 6 (histopathologically-confirmed malignancy) [12]. Herein, we were interested in evaluating the performance of LLMs in making a BI-RADS classification upon breast lesion ultrasound images.

To date, there have been few studies that have determined the performance of LLMs in making a classification using BIRADS. In a short communication article by Nguyen *et al*., 2025, it was found that GPT-4 and GPT-4o had a 66.2% accuracy among all breast cancer mammogram images [13]. The primary limitation of this study, like many studies evaluating LLMs, was that replicates were not performed to account for the inherent variability of responses provided from LLMs, and the possibility of the original image dataset being used for training these closed-source models [14]. Likewise, in another retrospective study, it was found that GPT-4, GPT-3.5, and Bard had differences between radiologists in classification, and would negatively impact clinical management of cases [15]. Furthermore, another study evaluating GPT-4 found that the model was frequently incorrect and had poor reproducibility [16]. Yet, to our knowledge, there has not been any comprehensive studies that have accounted for prompt engineering techniques, replicates, and large sample sizes of images for determining the performance of these models. Here, we evaluated the performance of Gemini 2.0 in evaluating 256 breast ultrasound lesions with account of prompt engineering techniques to effectively gauge the capabilities of the model.

## 2. Materials and Methods

### 2.1. Study Design

This study was conducted at the California University of Science and Medicine, School of Medicine, Department of Medical Education, Colton, California, USA. All data was collected from public sources, and therefore the presented study did not require approval from an Institutional Review Board or consent from patients. We accessed Google Gemini 2.0 Flash (https://gemini.google.com/) on February 10, 2025 to March 5, 2025. We did not tune or adjust the hyperparameters of the model, and only assessed its original performance. The ultrasound based breast lesion dataset, including 256 images from 256 different patients was obtained from Pawłowska et al., [17]. In the original article or the source webviewer server (https://best.ippt.pan.pl/datasets/breast), each image provides classification data including the age of the patient, tissue composition, symptoms, shape, margin, echogenicity, posterior features, halo, calcification, skin thickening, interpretation by radiologists, BI-RADS, verification, diagnosis. The demographics and the structure of the BrEaST dataset have been listed in the original publication. We opted to utilize this dataset over previously published models given the recency of the work, which has likely not been involved in training of the language model as of February 17, 2025.

### 2.2. Prompt engineering for the LLM

Given that the prompt used to identify the abilities of the LLM has a large influence on its accuracy and precision, we provided iterations of each individual patient image. For instance, the first iteration included: “Please act as an radiologist with extensive expertise in evaluating the breast. Tell me the BI-RADS score of this image. Be specific and include subtype if justified”. The second iteration is the following: You are a chief radiologist. Please tell me the BI-RADS score for this ultrasound of the breast. Be specific and include a subtype if justified.” Lastly, the third iteration of the prompt included: “You are a radiologist with expertise in breast imaging. Please evaluate this image comprehensively and provide me with the BI-RADS score of the following image. Be specific and include a subtype if justified.” To ensure that the model does not recall prior responses or patterns in the dataset, we inputted each iteration into a different context window (i.e. New chat). Furthermore, Gemini activity was turned off to ensure that the images provided to the models in the different context windows are not trained during the experimental period. As a final step to confirm that the model does not recall prior steps, we classified each image and randomized the order for which the images were displayed to Gemini.

### 2.3. Statistical Analyses

All statistical analyses were performed in GraphPad Prism (version 10.0.2). Depending on the context, we either assumed the data was normally distributed or not. For categorical data, we performed Fisher’s exact test. For multiple continuous variables, an ordinary one-way ANOVA was performed followed by Tukey’s post-hoc test if significant. The assumptions used for this study include α = 0.05, where a *p*-value below 0.05 was considered to be statistically significant. Depending on the context, a Student’s *t*-test or Wilcoxon ranked-sum test was performed. A power analysis was not conducted in this study due to the sample sizes either being low or above the CLT threshold, where it was assumed that only large significant differences observed are likely to be confirmed.

## 3. Results

The performance of Gemini 2.0 Flash in classifying the 256 breast lesion ultrasound images and its replicates has been visualized in **Fig. 1**, where the distribution of the ground truths and replicates, accuracy overall and of each category, and ROC-AUC analyses have been shown. We found that the distribution of predictions was significantly different between all groups (*p* < 0.0001), and most notably an outlier was observed in the second replicate making 137 predictions of the 4b category in 256 cases. An interesting limitation of the model was that it refused to give a response to the prompt or insisted that the image itself was not an ultrasound of the breast despite being told in the initial prompt in all replicates, which likely is a cause for the distribution of predictions being significantly different from the ground truth. The accuracies are: category 1 was 20% ± 11.55%, 2 was 11.11% ± 2.94%, 3 was 18.1% ± 5.79%, 4a was 4.44% ± 2.57%, 4b was 36.23% ± 10.82%, 4c was 22.92% ± 3.18%, 5 was 17.73% ± 5.54%, and the overall accuracy of the model was 19.01% ± 1.63%. Given the accuracy metric does not consider the correct predictions of true negative results, we calculated the receiver operating characteristic-area under the curve (ROC-AUC) analyses. Given that the ROC-AUC analyses require binary classification, we performed the analyses on individual categories. For instance, with the ROC-AUC analyses of category 4c for the first replicate iteration, ultrasound images with classification of 4c were observed to be true positive while any other category predictions would be considered false positives. Our analyses were conducted on all 7 categories from the 3 iterations, culminating in 21 ROC curves provided in the **Supplementary Data**. We then took the AUC, SEM, and sample size (*n* = 256) per replicate and summarized the data in a forest plot (**Fig. 1C**). The AUC of category 1 was 0.5078 ± 0.0013, 2 was 0.5319 ± 0.0064, 3 was 0.5293 ± 0.0039, 4a was 0.5553 ± 0.0073, 4b was 0.5729 ± 0.05347, 4c was 0.5247 ± 0.0101, and 5 was 0.5495 ± 0.0169, respectively. While there was no significant differences in AUC between the different categories (*p* < 0.0001), categories 1 (*p* = 0.02), 2 (*p* = 0.0382), 3 (*p* = 0.0173), 4a (*p* = 0.0168) were significantly greater than the 0.5 benchmark and 4b (*p* = 0.3058), 4c (*p* = 0.134), and 5 (*p* = 0.1001) were not. These results suggest that Gemini 2.0 performed significantly better at identifying benign or with a low probability of malignancy compared to those with higher probabilities. Still, the performance of the model (AUC = 0.5 - 0.6) was low compared to those of the radiologists.

**Figure 1.**
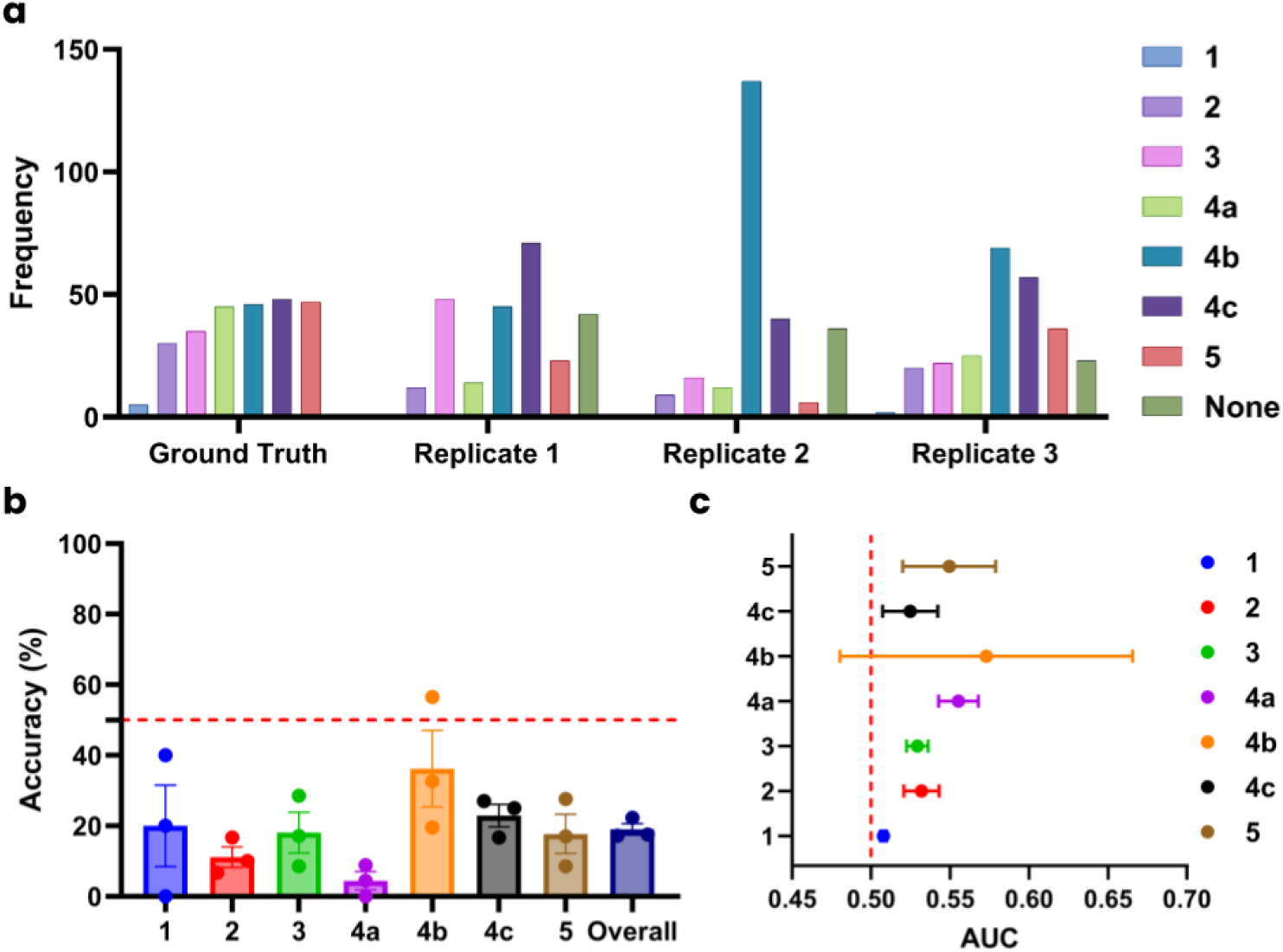
Performance of Gemini in BI-RADS classification of 256 breast lesion ultrasound images. (a) Distribution of BI-RADS classifications for the ground truth and iterations evaluated in this study. (b) Accuracy of the model for each category and overall dataset (total correct/sample size). Data displayed as mean ± SEM. (c) Forest plot of ROC-AUC analyses for each category with benchmark at AUC = 0.5. Data displayed as mean ± SD for better visualization of the AUC benchmark.

Given the results from the statistical and ROC-AUC analyses, we conclusively expressed interest in determining the closeness of incorrect predictions to the ground truth. We assigned each category a numerical value (i.e. ‘1’ is 1,… ‘4a’ is 4, ‘4b’ is 5, ‘4c’ is 6) excluding predictions of ‘none’. The category differences were calculated by subtracting the predicted value from Gemini by the ground truth, followed by taking the absolute value of the difference. In **Fig. 2**, we have visualized the distribution of predictions compared to the true positive category as bar graphs for each replicate to distinctly highlight deviation among the distributions and individual predictions provided from the model. Category difference was calculated by the value. From one-sample *t*-tests of each category, we generally found that all incorrect predictions 1-2 categories away from the ground truth (*p*-values provided in **Supplementary Data**). These findings are critical to defining the precision of LLMs from achieving the accurate prediction. Overall, these results suggest that Gemini lacks the precision necessary to be useful in disease diagnostics, and further training is necessary before realistic clinical usage.

**Figure 2.**
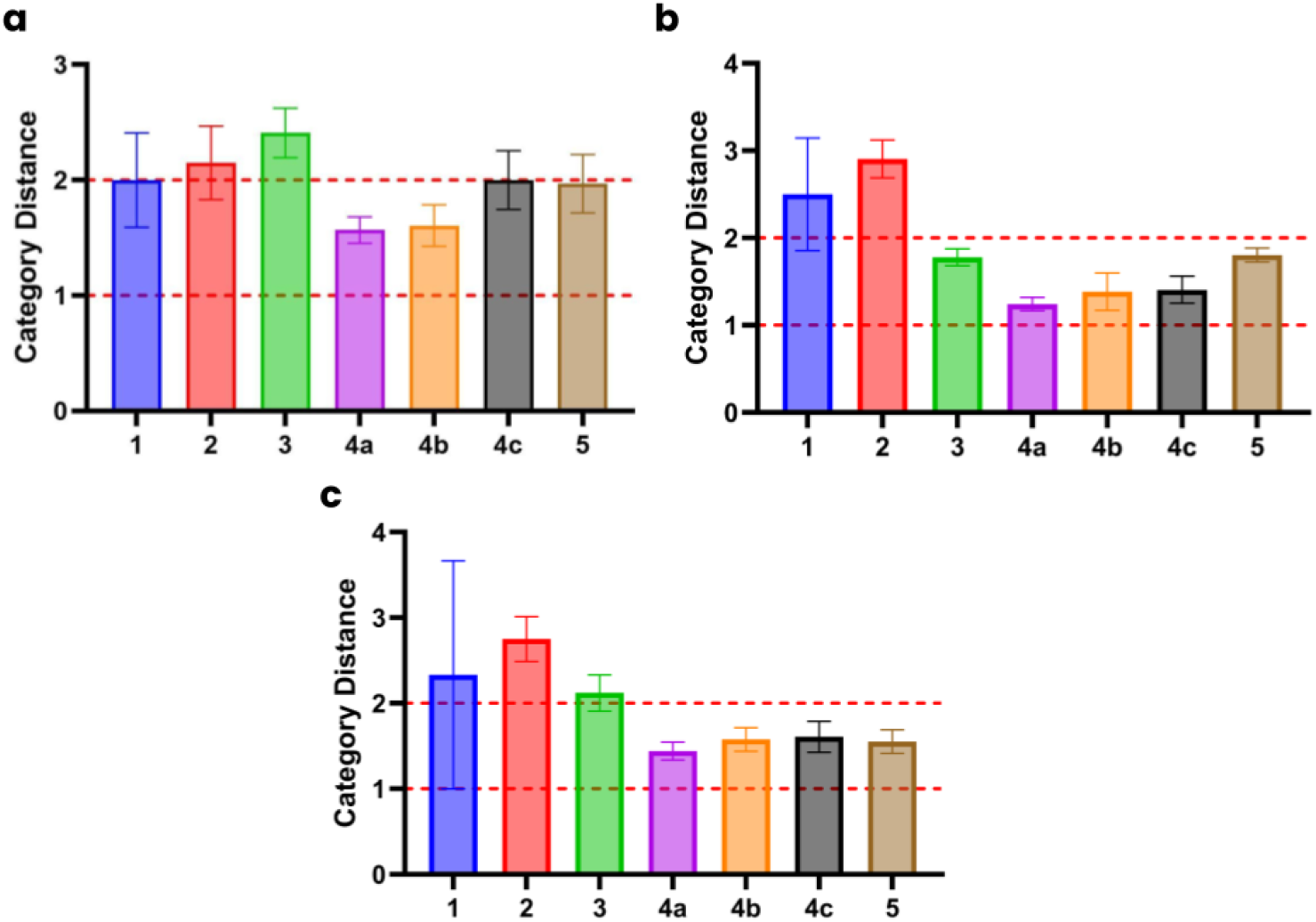
Distribution of differences in category. The distance is calculated by the difference between the prediction and the ground truth for each category displayed in the **(a)** first, **(b)** second, **(c)** third replicate. All data has been displayed as the mean ± SEM.

## 4. Discussion

To our knowledge, this is the first comprehensive cross-sectional study to benchmark the performance of LLMs in making BI-RADS classifications with replicates. We used a large breast ultrasound dataset including 256 unique lesions that have received complete labeling by radiologists to evaluate the Gemini 2.0 model from Google. This study found that the model had an accuracy of 19%, capable of deciphering benign lesions while those malignant were akin to chance. Overall, this work emphasizes the current limitations of LLMs and their potential application in clinical imaging. Further training is required for multimodal language models in the ultimate goal of understanding, processing, and improving clinical decision-making before realistic healthcare integration.

Our study is also supported by multiple similar studies reported in the past year. As previously described, Nguyen *et al*., [13] found that GPT-4 and GPT-3.5 had a 66% accuracy similar to Haver *et al*., [16]. Recently, though, Güneş *et al*., [18] benchmarked ChatGPT (4o, 4, and 3.5), Gemini Pro 1.5, and other publicly-available models on 100 multiple choice questions from the American College of Radiology and 100 ultrasound breast images. The authors found that Gemini 1.5 Pro had an accuracy of 61% in text-based questions but 31% for classifying BI-RADS scores for images. Güneş *et al*., [18] directly supports the conclusions of this study, and further suggests that other commercial models have a stronger performance in making classifications. Overall, the key message of this work is that further studies and training are required for the vision feature of multimodal LLMs before realistic usage in BI-RADS classification.

## Data Availability

Source data is available at Pawłowska et al., (2024). The statistical analyses performed in this study have been provided in the Supplemental Material.

## Supplementary Material

The Ultasound_Data.prism file has been provided for all visualizations and analyses performed in this study.

## Declaration of Conflicts of Interest

All authors declare no conflicts of interest.

## Acknowledgements

None to report.

## Notes

**CONFLICTS OF INTEREST:** The authors have no competing interests.

### Competing Interest Statement

The authors have declared no competing interest.

### Funding Statement

None to declare.

### Author Declarations

The human breast ultrasound images used in this study are publicly available from Pawłowska et al., (2024).

